# Human Germline Heterozygous Gain-of-Function *STAT6* Variants Cause Severe Allergic Disease

**DOI:** 10.1101/2022.04.25.22274265

**Authors:** Mehul Sharma, Henry Y. Lu, Maryam Vaseghi-Shanjani, Kate L. Del Bel, Oriol Fornes, Robin van der Lee, Phillip A. Richmond, Susan Lin, Joshua Dalmann, Jessica J. Lee, Allison Matthews, Géraldine Blanchard-Rohner, Clara D M van Karnebeek, H. Melanie Bedford, Wyeth W. Wasserman, Michael Seear, Margaret L. McKinnon, Hanan Ahmed, Stuart E. Turvey

## Abstract

STAT6 (Signal transducer and activator of transcription 6) is a transcription factor that plays a central role in the pathophysiology of allergic inflammation. STAT6 mediates the biological effects of IL-4, a cytokine necessary for type 2 differentiation of T cells and B cell survival, proliferation and class switching to IgE. We have identificated two unrelated patients with a phenotype notable for their early-life onset of profound allergic immune dysregulation, widespread treatment-resistant atopic dermatitis, hypereosinophilia with esosinophilic esophagitis, elevated serum IgE, IgE-mediated food allergies, and vascular anomalies of the brain. Both patients harbored heterozygous *de novo* missense variants in the DNA binding domain of *STAT6* (c.1144G>C, p.E382Q; and c.1256A>G, p.D419G). Functional studies established that both variants caused a gain-of-function (GOF) phenotype associated with enhanced phosphorylation and transcriptional activity of STAT6, in addition to increased transcript abundance of known STAT6 target genes and other genes implicated in allergic disease. JAK inhibitors decreased the enhanced STAT6 responses associated with both these STAT6 GOF variants. This study identifies heterozygous GOF variants in *STAT6* as a novel autosomal dominant allergic disorder. We anticipate that our discovery of the first humans with germline *STAT6* GOF variants will facilitate the recognition of more affected individuals and the full definition of this new primary atopic disorder.

## INTRODUCTION

Signal transducer and activator of transcription (STAT) proteins are a family of seven mammalian transcription factors (STAT1, STAT2, STAT3, STAT4, STAT5A/B, and STAT6) responsible for regulating the expression of genes involved in cellular immunity, survival, proliferation, and differentiation^1,2^. Although they each have unique functions, they share sequence homology and possess similar activation mechanisms^3^. In general, upon extracellular cytokine/growth factor-receptor engagement, intracellular membrane receptor-associated Janus kinases (JAK) undergo transphosphorylation and activation, leading to receptor tyrosine phosphorylation^4^. Phosphorylated tyrosine residues on the receptor then create binding sites for the normally cytosolic-located Src homology 2 (SH2) domain-containing STATs to bind and become phosphorylated and activated. Phosphorylated STATs, dimerize and translocate into the nucleus, where they bind to specific DNA sequences and regulate transcription^4-7^. As such, STATs are critical for transducing extracellular cytokine-induced responses to transcriptional changes in the nucleus.

Signal transducer and activator of transcription 6 (STAT6) is the main transcription factor that mediates the biological effects of IL-4, a cytokine necessary for type 2 differentiation of T cells, as well as B cell survival, proliferation and class switching to IgE^8-11^. IL-4 is predominantly secreted by T cells, mast cells, basophils, and type 2 innate lymphoid cells^12^. To mediate its effects, IL-4 binds the widely expressed IL-4Rα and this complex then binds secondary receptors chains^13^. In hematopoietic cells, the secondary receptor chain is the common γc chain also used by IL-2, IL-7, IL-9 and IL-15 and this forms the type 1 IL-4R system^14^. In non-hematopoietic cells such as epithelial cells, the secondary receptor chain is IL-13Rα, which forms the type 2 IL-4R system^13^. Activation of these two receptor systems lead to diverse functions. In B cells, IL-4-STAT6 signalling supports the growth and differentiation of B cells as well as secretion of pro-allergic immunoglobulin E (IgE)^9^. In T cells, the IL-4-STAT6 axis is critical for the differentiation of T helper 2 (T_H_2) cells^15^, a subset of CD4^+^ helper T cells that is a major contributor to the pathogenesis of allergic disease. In epithelial cells, the IL-4/IL-13-STAT6 pathway has been implicated in barrier dysfunction^16,17^ and chronic pruritus^18^. Collectively, the IL-4-STAT6 axis drives the development and progression of allergic disease and asthma. Indeed, a monoclonal antibody directed against IL-4Rα (dupilumab) is clinically approved for treating allergic inflammation^19,20^. Mouse studies further emphasize the central role that STAT6 plays in the allergic diathesis. Mice lacking STAT6 fail to develop many of the features of allergic inflammation^21-23^.

In 2018, the term primary atopic disorders (PADs) was coined to describe heritable genetic disorders presenting with dysregulated pathogenic allergic effector responses^24-26^. PADs are a subgroup of inborn errors of immunity (IEIs) that manifest with prominent allergic inflammation, in the presence or absence of other clinical features associated with IEIs, such as enhanced susceptibility to infections, autoimmunity, and malignancy. Of the ∼40 monogenic causes of PADs described to date, 12 impact the JAK-STAT signaling cascade that is critical for cytokine responsiveness. For example, both loss-of-function (LOF) and gain-of-function (GOF) variants in *STAT3*^27,28^ and *STAT5B*^29,30^, and GOF variants in *JAK1*^31,32^ and *STAT1*^33^ cause PADs. Furthermore, defects in the genes encoding upstream and downstream proteins in the STAT pathway (e.g. *ZNF341, DOCK8, IL6ST, IL6R, and ERBIN)* also result in severe allergic diseases^26^. The recent increased recognition of PADs is valuable because: (i) the identification of affected pathways provide important insights into the pathogenesis of allergic inflammation and may uncover new treatment targets; (ii) these conditions are likely under-diagnosed since allergies are traditionally believed to be complex polygenic disorders; and (iii) diagnosis of PADs can be transformative for affected individuals and their families by guiding treatment options, and informing relevant considerations tied to genetic counselling including family planning, long term prognosis, and connection to other families with the same genetic disease.

In this study, we describe a novel human PAD caused by germline heterozygous GOF variants in the DNA binding domain of *STAT6* found in two unrelated individuals with severe allergic disease and vascular anomalies of the brain.

## METHODS

### Ethical Considerations

All study participants provided written informed consent. Research study protocols were approved by The University of British Columbia Clinical Research Ethics Board.

### Identification of *STAT6* variant via whole exome sequencing

Trio whole exome sequencing (WES) was performed as previously described^34^. Briefly, WES was performed on genomic DNA extracted from the first patient (P1) and both his parents and a novel *de novo* predicted damaging *STAT6* variant (c.1144G>C p.E382Q) was selected for further analysis. WES was also performed on genomic DNA extracted from the second patient (P2), her mother, and one of her brothers. A novel *de novo* predicted damaging variant for *STAT6* (c.1256A>G, p.D419G) was identified. Sanger sequencing on saliva samples collected from the remaining family members of P2 revealed that no other individuals carried the variant and that it segregates with disease.

### Generation of *STAT6* variant plasmids

Plasmids used for transfection studies contained full-length *STAT6* in a pCMV6 entry vector with a C-terminal GFP tag (Cat#: RG210065, OriGene Technologies; Rockville, Maryland, USA). To generate p.E382Q and p.D419G STAT6, a Q5 site-directed mutagenesis kit (Cat# E0554S, New England Biolabs, Ipswich, MA, USA) was used according to manufacturer’s recommendations along with the following primer pairs: 5’-TGTCACAGAG<u>C</u>AGAAGTGCGC-3’, 5’-GACTCAGTGCCCTTCCGC-3’, and 5’-GGCAACCAAG<u>G</u>CAACAATGCC-3’, 5’-ATGGACGATGACCACCAG-3’, respectively.

To generate lentivirus vectors, wild type (WT), p.E382Q, and p.D419G STAT6 from the above plasmids were cloned into a GFP-tagged Lenti vector (Cat#: PS100071, OriGene Technologies, Rockville, MD, United States) using EcoRI-HF (Cat#: R3101) and NotI-HF (Cat#: R3189) both from New England BioLabs. The three STAT6 plasmids were packaged using 3rd generation packaging plasmids and transfected into HEK293T cells. Culture media was collected, centrifuged, filtered, concentrated, and stored at - 80°C before use.

All new variant plasmids were confirmed by Sanger sequencing and purified from 10-beta competent *E. coli* using a QIAprep Spin Miniprep Kit (Qiagen, ON, Canada).

### Transient and stable expression of *STAT6* variants

Transient expression of *STAT6* variants in HEK293 cells were accomplished using a Lipofectamine 3000 kit (Thermo Fisher Scientific, Waltham, MA, USA) according to manufacturer’s recommendations. Briefly, HEK293 cells were seeded at 8.0 × 10^5^ cells/well in a 6-well plate in 1.5 mL of Dulbecco modified Eagle medium (DMEM) with 10% FBS (Gibco, Life Technologies; Rockville, MD, USA) and incubated for 24 hours at 37°C. Cells were transfected with 2.5μg of plasmid DNA using the P3000 and Lipofectamine 3000 reagents and harvested after 24h.

Stable expression of STAT6 in Jurkat T cells was accomplished using the Lentivirus approach as previously described^35,36^. Briefly, Jurkat T cells were infected with lentiviral particles in the presence of 5μg/ml polybrene (Sigma-Aldrich, St Louis, MO, USA) and spinoculated at 800 x g for 30mins at 32°C, cultured, and expanded in complete RPMI-1640 (GE Healthcare, Chicago, IL, United States) supplemented with 10% FBS. Expanded cells were sorted on GFP expression using a BD FACS Aria (BD Biosciences) cell sorter.

### Determining *STAT6* variant activity by phospho-flow cytometry

Phospho-STAT6 activity was quantified by flow cytometry in transfected cells stimulated with 100ng/mL of IL-4 (Cat# 204-IL-020, R & D Systems, Minneapolis, MN, USA) for 1h or transfected cells pre-treated with either 10μM ruxolitinib (Cat code: tlrl-rux, Invivogen, San Diego, CA, USA) or 4μM tofacitinib (Cat# S50001, Selleckchem, Houston, TX, USA) for 2h before stimulation as previously described^37^. Briefly, transfected and stimulated cells were fixed using BD Cytofix (Cat# 554655, BD Biosciences, Ontario, Canada) for 20 min at 4°C and permeabilized using Perm III for 30 min on ice (Cat# 558050 BD Biosciences). The cells were then stained with STAT6 PE-CF594 (Cat# 564148, BD Biosciences) and p-STAT6 AF647 (Cat# 612601, BD Biosciences). pSTAT6 expression was measured in GFP^+^STAT6^+^ cells from WT and *STAT6* variants samples on an LSRII flow cytometer (BD Biosciences) and analyzed using FlowJo software (BD Biosciences).

### Luciferase reporter assays

A luciferase reporter plasmid encoding a 4x STAT6 binding site [<u>TTC</u>CCAA<u>GAA</u>] was used to assess WT and variant STAT6 promoter activity^38^. The p4xSTAT6-Luc2P plasmid was a gift from Axel Nohturfft (Addgene plasmid #35554; http://n2t.net/addgene:35554; RRID:Addgene_35554). Briefly, HEK293 cells were seeded in 24-well plates overnight at a density of 150k cells and transfected with 250ng of p4xSTAT6-Luc2P, 250ng of a plasmid encoding GFP-tagged WT, p.E382Q, or p.D419G *STAT6*, and 10ng Renilla luciferase (R-Luc) using lipofectamine 3000 as described above. Transfected cells were subsequently stimulated with 100ng/mL of IL-4 (Cat# 6507-IL-010, R&D Systems, Minneapolis, MN, USA) or left untreated. Cell lysates were prepared and processed using a Dual-Glo Luciferase Assay Kit (Cat#: E2920, Promega, Madison, WI, USA) according to manufacturer’s recommendations. Luciferase activity was measured on the Infinite M200 plate reader (Tecan; Männedorf, Switzerland).

### RNA sequencing

To investigate the global transcriptome, *STAT6* variant-transduced Jurkat T cells were left unstimulated, or stimulated with 100ng/mL IL-4 for 4h. RNA was extracted in triplicate as previously described^37^ using a RNeasy Mini Plus Kit (Qiagen) according to manufacturer’s recommendations. RNA was prepared following the standard protocol for the NEBNext Ultra II Stranded mRNA (New England Biolabs) and sequenced on the Illumina NextSeq 500 with Paired End 42 bp × 42 bp reads. De-multiplexed read sequences were aligned to a reference sequence using RNA-Seq Alignment app (v1.1.1) on Illumina Basespace, using Spliced Transcripts Alignment to a Reference (STAR) aligner and Cufflinks 2 for assembly and estimation of gene expression.

Expression data were normalized to reads between samples using the edgeR package in R (R Foundation, Vienna, Austria). Normalized counts were filtered to remove low counts using the filterByExpr function in edgeR^39^. Principal component analysis (PCA) was done on log_2_(normalized counts+0.25) in R using the PCA function. Differential expression between unstimulated and stimulated samples for all three *STAT6*-transduced Jurkat T cells was accomplished using Limma^40^. Differentially expressed genes were defined as those with fold change (FC) greater than 1.25 and adjusted p-value less than 0.05.

Pathway analysis was done by first performing Gene Set Enrichment Analysis (GSEA) with 1000 permutations using the Molecular Signatures Database Hallmark module. Signal-to-noise ratio was used for gene ranking and the obtained NESs and P-values were further adjusted using the Benjamini-Hochberg method. Pathways with an adjusted P-value < 0.05 were considered significant. Leading edge genes from significant pathways between *STAT6* WT and *STAT6* c.1256A>G Jurkat were identified (no significantly upregulated pathways were identified between *STAT6* Wild Type and *STAT6* c.1144G>C). Expression levels of these genes were then determined in each of the three groups (Wild Type, c.1144G>C, c.1256A>G) under both stimulated and unstimulated conditions. Sample level enrichment analyses (SLEA) scores were computed as previously described^41^. Briefly, z-scores were computed for gene sets of interest for each sample. The mean expression levels of significant genes were compared to the expression of 1000 random gene sets of the same size. The difference between observed and expected mean expression was then calculated and represented on heatmaps.

Gene set enrichment analysis was also done on two other gene sets: i) STAT6 targets; and ii) IL-4 T_H_2 targets. STAT6 targets were defined as the set of genes that were significantly upregulated upon IL-4 stimulation in WT-STAT6 transduced Jurkat T cells. Enrichment of this gene set was determined at both baseline and after IL-4 stimulation between WT and p.E382Q STAT6, and WT and p.D419G STAT6 Jurkat T cells. IL-4-STAT6 T_H_2 skewing targets were previously reported^42^ STAT6 target genes that are significantly upregulated in response to IL-4 treatment and that lead to T_H_2 skewing.

Raw data will be deposited on Gene Expression Omnibus.

### Structural modeling

The effects of the STAT6 variants on the protein function and structure were analyzed using three-dimensional models. We used SWISS-MODEL^43^ to model the variants based on a template structure of the human STAT6 transcription factor solved as a homodimer and in complex with DNA (PDB: 4Y5W, resolution: 3.1 Å, chains A, C, M and N)^38^. Structures were visualized with UCSF Chimera^44^.

## RESULTS

### Clinical features of two patients with severe atopic disease

Patient A-II-1 (P1) was a male patient who had life-long severe allergic disease. The patient was not dysmorphic and was born to unrelated parents. Soon after birth, P1 developed widespread, treatment-resistant atopic dermatitis (eczema), gastroesophageal reflux with episodes of aspiration pneumonia, as well as chronic diarrhea. He underwent Nissen fundoplication a few months later (with a surgical revision in early childhood, age 6-10 years), following which gastrostomy feeding was commenced. Over time, he was also diagnosed with multiple food and drug allergies with anaphylactic reactions (combined with positive skin prick testing or RAST testing) documented in response to peanuts, cows milk protein, bananas, sesame seeds, fish, and eggs. He was fed with extensively hydrolyzed and elemental formulas via gastrostomy with very limited oral feeding for much of his life. He was also diagnosed with asthma. He experienced episodes of secondary bacterial skin infections from which *Staphylococcus aureus*, group A streptococci and *Candida* spp. were isolated. Widespread *molluscum contagiosum* was another notable skin finding. In his second and third decade of life, he had persisting eczema, asthma, and perennial rhinitis which worsened during pollen seasons. Repeated upper GI endoscopies confirmed the diagnosis of eosinophilic esophagitis. During this time, most of his feeding consisted of elemental formula via gastrostomy supplemented with limited oral intake (mainly rice, potatoes, and chicken). He was treated with oral steroids for many years and experienced symptom flares whenever the dose of prednisone was dropped below 10mg/day. He developed side-effects related to corticosteroids including cataracts, osteoporosis, and pathogenic fractures. He had an episode of thromboembolic ischemia of the right first and third toes with pain and cyanosis, ultimately requiring a lumbar sympathectomy to relieve the pain. In his mid-30s (age 30-35 years), he was hospitalized for non-bloody diarrhea, profound weight loss, malnutrition, weakness, and vertigo. He was treated with high dose of prednisone, antibiotics, and elemental feeding. During the hospitalization, he died after developing a spontaneous subarachnoid hemorrhage with obstructive hydrocephalus, presumed secondary to an aneurysm (**Table 1**). Blood testing over the years confirmed eosinophilia and high serum IgE levels (**Fig 1**). Lymphocyte phenotyping and proliferation to a panel of mitogens were found to be unremarkable when measured in early adulthood (ages 20-25 years). An initial genetic assessment revealed normal sequences for *STAT3* and *DOCK8*, prompting a more extensive genetic evaluation by WES.

**Table 1:** Major Clinical Features. Tabulation and comparison of the clinical phenotype of patient 1 and 2.

|  | Patient 1 | Patient 2 |
| --- | --- | --- |
| **Genetic Workup** | Heterozygous *STAT6*<br>c.1144G>C, p.E382Q | Heterozygous *STAT6*<br>c.1256A>G, p.D419G |
| **Severe Atopic Disease** |  |  |
| Disease onset in early infancy | ✓ | ✓ |
| Severe, widespread treatment-resistant atopic dermatitis | ✓ | ✓ |
| Asthma | ✓ | ✓ |
| Multiple food allergies | ✓ | ✓ |
| Drug allergies | ✓ | ✓ |
| Allergic rhinoconjunctivitis | ✓ | ✓ |
| **Infections** |  |  |
| Recurrent respiratory infections | ✓ | ✓ |
| Recurrent skin infections (e.g. *Staphylococcus*, *Candida*, *molluscum*) | ✓ | ✓ |
| **Gastrointestinal Disorders** |  |  |
| Early gastro-oesophageal reflux disease (GERD) | ✓ |  |
| Eosinophilic esophagitis |  | ✓ |
| Chronic diarrhea, protein losing enteropathy, eosinophilic infiltration, duodenal atrophy | ✓ |  |
| **Skeletal Issues** |  |  |
| Osteoporosis, pathogenic fractures | ✓ |  |
| Generalized hypermobility and joint pain |  | ✓ |
| **Vascular anomalies** |  |  |
|  | In late 20s:<br>Thromboembolic ischemia of 1st and 3rd toes with severe pain | Brain MRI in mid-30s (age 30-35 years) revealed multiple anatomical variants including anomalous vasculature in the circle of Willis with hypoplastic vertebrobasilar arteries, a persistent left-sided congenital trigeminal artery with tortuosity, and a hypoplastic A1 segment of the right anterior carotid artery. |
|  | Mid-30s (age 30-35 years): Fatal spontaneous subarachnoid hemorrhage with intraventricular hemorrhage. |  |
| Other |  | Adolescence (age 15-20 years): Retinal detachment in right eye |

**Figure 1:**
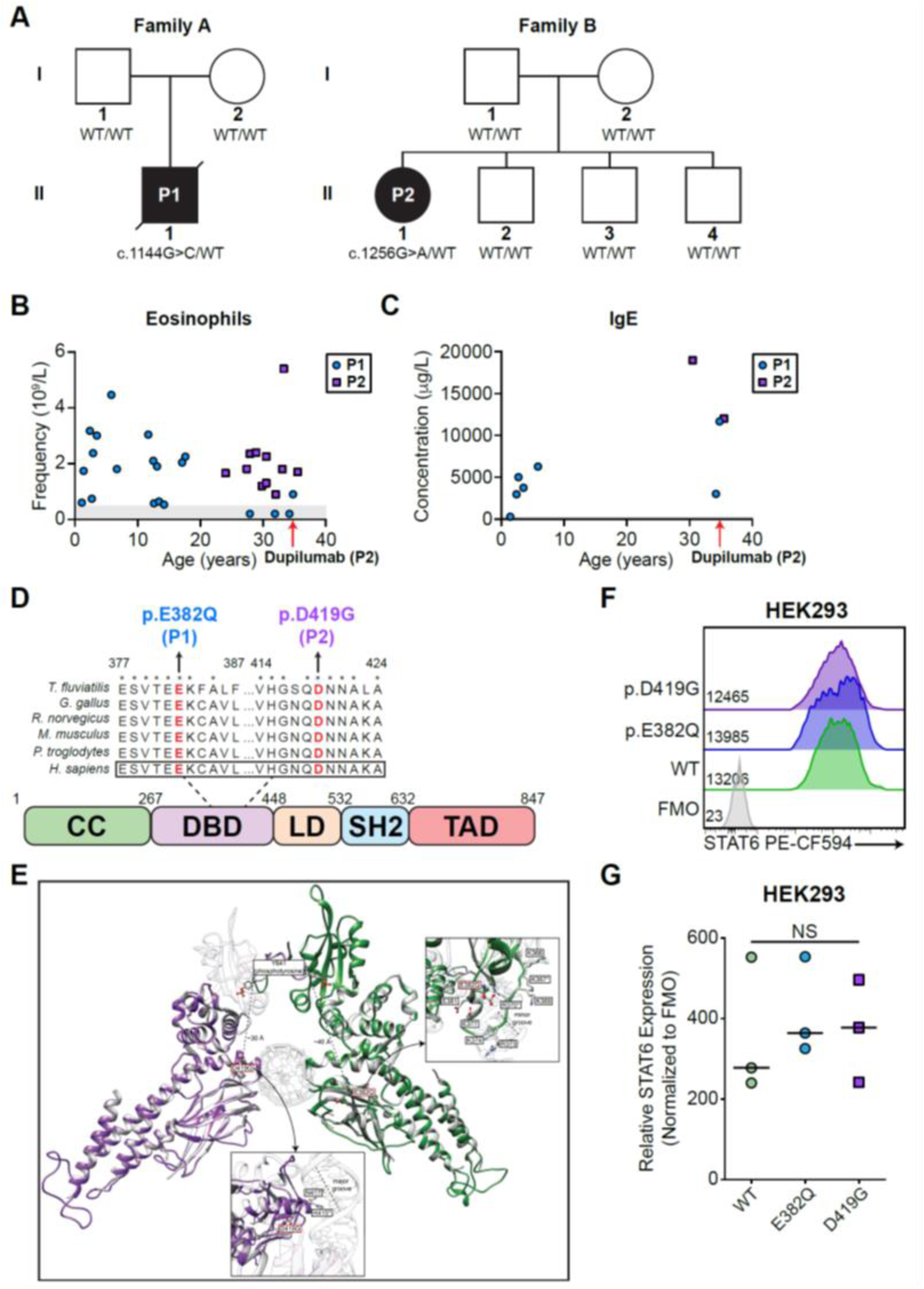
Clinical phenotype of two patients with severe atopic disease. **A)** Family pedigree of the two patients. Filled symbols=affected individual, arrow=proband, unfilled symbols=unaffected individual. P1 has the variant p.E382Q, P2 has the variant p.D419G. **B)** Eosinophil frequency in whole blood of P1 and P2. **C)** IgE concentration in whole blood of P1 and P2. **D)** Schematic illustrating the protein domains of STAT6. Amino acid location of the variants shown in red. Affected region was aligned to other species. Asterisks indicate full conservation. **E)** Structural model of the DNA-STAT6 homodimer complex. Green=p.E382, Purple=p.D419. Residues marked with an asterisk (*) were demonstrated to affect the binding of STAT6 to DNA in a previous study^38^. **F)** STAT6 expression in WT-, p.E382Q-, and p.D419G-*STAT6*-transfected HEK293 cells. **G)** Quantification of relative protein expression in F), n=3. Significance determined through a one-way ANOVA and Tukey’s post-hoc test.

Patient B-II-1 (P2) is a female who had similar life-long struggles with severe allergic disease. She presented with severe chronic, treatment-resistant atopic dermatitis since birth. She was diagnosed with IgE-mediated food allergy to egg, milk, peanut and tree nuts early in life (age 0-5 years). She was diagnosed with asthma as an infant. In her early life (age 0-5 years) she also had a pneumothorax requiring chest tube insertion, required intubation for an asthma exacerbation, and was diagnosed with giant papillary conjunctivitis which has remained an ongoing challenge. Other eye manifestations included unilateral retinal detachment in her adolescence (age 15-20 years) and the development of cataracts requiring surgery in early adulthood (age 20-25 years). Through her second and third decades of life she continued to suffer from eczema, asthma, recurrent bronchitis, rhinoconjunctivitis, and secondary staphylococcal and candida skin infections. She had multiple dental abscess in the second and third decades of her life. In her 20s she began experiencing gastrointestinal symptoms culminating in biopsy diagnosed eosinophilic esophagitis and eosinophilic gastroenteritis. Joint examination confirmed the presence of hypermobility. Serial blood testing over the years confirmed eosinophilia and high serum IgE levels (**Fig 1**). Brain MRI in her mid-30s (ages 30-35 years) revealed multiple anatomical variants including anomalous vasculature in the circle of Willis with hypoplastic vertebrobasilar arteries, a persistent left-sided congenital trigeminal artery with tortuosity, and a hypoplastic A1 segment of the right anterior carotid artery (**Table 1**). Multiple therapies were trialed over the years with corticosteroids offering the most obvious benefits. To seek a diagnosis for this spectrum of severe allergic manifestations, WES was performed on the patient, her mother and one of her brothers.

### Whole-exome sequencing reveals novel *de novo* heterozygous variants in the DNA binding domain of *STAT6*

The patients were found to carry heterozygous variants in the gene *STAT6* (NM_001178079.2): c.1144G>C, p.E382Q for P1 and c.1256A>G, p.D419G for P2. The variants were *de novo* as they were not found in any other family members (**Fig 1A**). Both variants have not been reported in population databases (i.e. gnomAD), but p.D419G can be found in the Catalogue of Somatic Mutations in Cancer (COSMIC) database as a recurrent somatic variant in lymphoma with experimental evidence of a GOF phenotype for this variant^45,46^. Pathogenicity prediction models predict both variants to be pathogenic, including CADD (Version 1.6, p.D419G: 29.3, p.E382Q: 27.7)^47^, SIFT (p.D419G: 0.001, p.E382Q: 0.026)^48^, and Polyphen-2 (p.D419G: 1.0, p.E382Q: 1.0)^49^ (**Supp Table 1**). Clinical laboratory tests showed that both patients had eosinophilia and markedly elevated IgE (**Fig 1B-C**). Both variants localized to the highly conserved DNA binding domain of STAT6 (**Fig 1D**). Glu382 and Asp419 lie close to the regions responsible for protein-DNA interaction^38^. Both of these variants decrease the electro-negativity of the protein near the DNA-binding interface (due to loss of glutamic acid (E) and aspartic acid (D)), and are predicted to enhance STAT6 binding to DNA (**Fig 1E**). These variants did not affect STAT6 protein expression as measured by flow cytometry in transfected HEK293 cells (**Fig 1F-G**).

### p.E382Q and p.D419G *STAT6* variants lead to increased phosphorylation and transcription activity of STAT6 in HEK293 cells

To assess the functional impact of the p.E382Q and p.D419G STAT6 variants, we selected HEK293 cells as our system, as these cells lack endogenous STAT6 but express other components of the IL-4R pathway (**Fig 2A**)^50^. HEK293 cells were transfected with wild-type (WT), p.E382Q, or p.D419G STAT6 and the phosphorylation status of STAT6 was quantified by flow cytometry after 24h (**Fig 2B**). At baseline, we observed significantly increased phosphorylated STAT6 in cells expressing the p.E382Q and p.D419G variants when compared to WT. IL-4 stimulation led to increased phospho-STAT6 in all conditions, with the variant expressing cells maintaining significantly higher phospho-STAT6 than WT (**Fig 2C**). To investigate whether increased phosphorylation of STAT6 in both variants lead to increased STAT6 transcription factor activity, we conducted luciferase assays with a reporter plasmid containing a canonical 4x STAT6 binding site, TTCCCAAGAA in HEK293 cells^38^. Both p.E382Q and p.D419G STAT6 variants resulted in significantly higher promoter activity at baseline (**Fig 2D**). However, there was no significant difference in promoter activity after 4h IL-4 stimulation (**Fig 2D**). Together these data confirm a GOF phenotype caused by both p.E382Q and p.D419G STAT6 variants.

**Figure 2:**
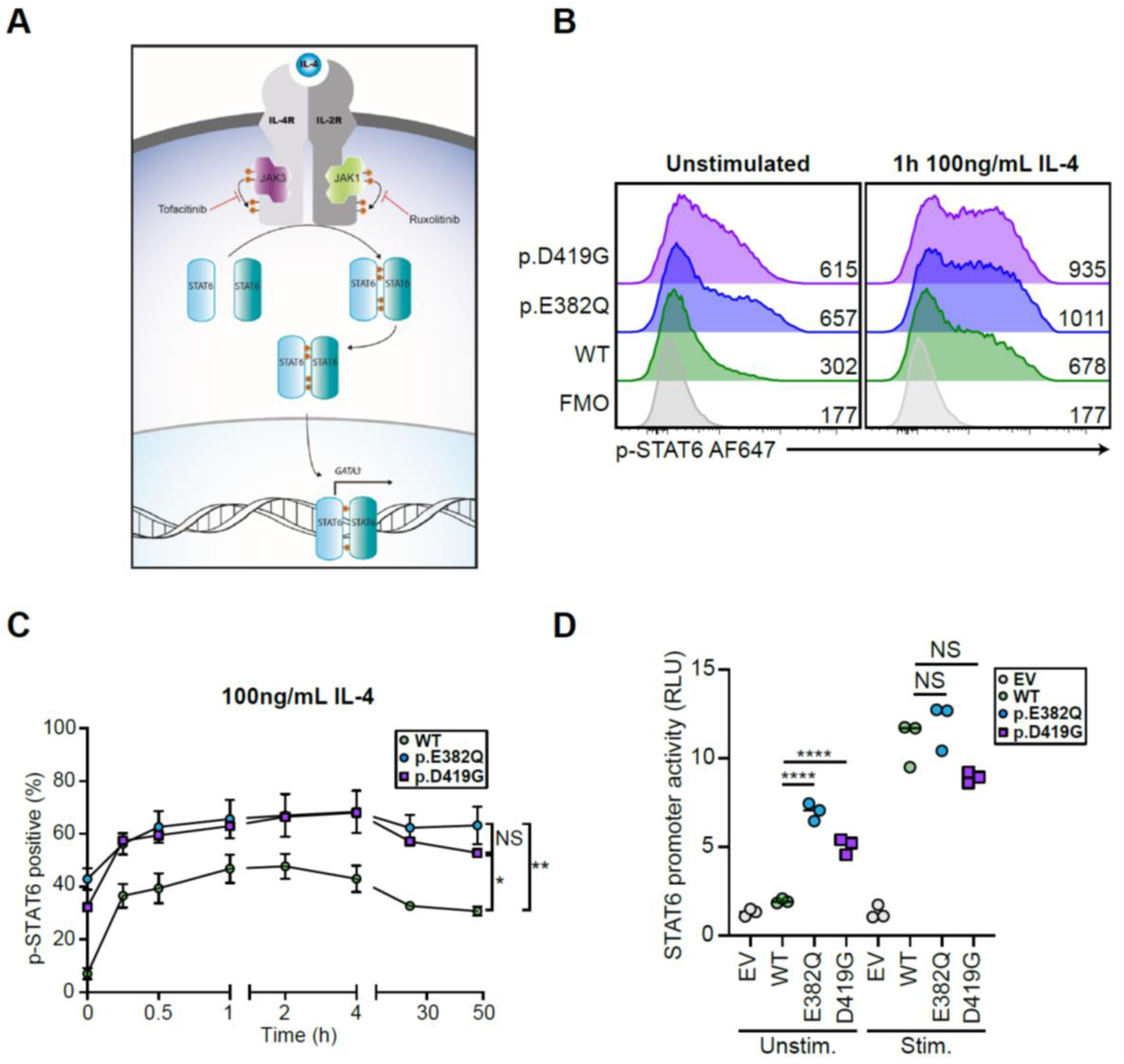
p.E382Q and p.D419G *STAT6* variants lead to increased STAT6 activity in HEK293 cells. A) Schematic illustrating classical IL-4-mediated STAT6 activation, dimerization, and phosphorylation. B) Phospho-STAT6 expression in WT-, p.E382Q-, and p.D419G-transfected HEK293 cells before and after treatment with IL-4 (100ng/mL for 1h). C) Time course of phospho-STAT6-positive cells after 100ng/mL IL-4 stimulation in transfected HEK293 cells, n=3. D) Luciferase assay of STAT6 activity on a plasmid containing a 4x STAT6 binding site [<u>TTC</u>CCAA<u>GAA</u>] for WT-, p.E382Q-, and p.D419G STAT6-transfected HEK293 cells before and after stimulation with IL-4 (100ng/mL for 4h), n=3. One-way ANOVA and Tukey’s post-hoc test. *p<0.05, **p<0.01, ***p<0.001, ****p<0.0001.

### p.E382Q and p.D419G STAT6 variants exert a gain-of-function phenotype in Jurkat T cells

Given the increased promoter activity of the p.E382Q and p.D419G STAT6 variants, we were interested in evaluating how this may impact the global transcriptome. As transcriptomic studies on HEK293 cells after IL-4 stimulation have been challenging to interpret^45^, we stably expressed p.E382Q and p.D419G STAT6 by lentivirus transduction in Jurkat T leukemia cells, which express endogenous STAT6 and serve as a model of heterozygosity^51^. Here, we observed significantly higher phospho-STAT6-positivity at baseline for the p.E382Q-cells but not p.D419G-cells, when compared to WT (**Fig 3A**). However, there was no significant difference in phospho-STAT6 positivity after IL-4 stimulation (**Fig 3A**).

**Figure 3:**
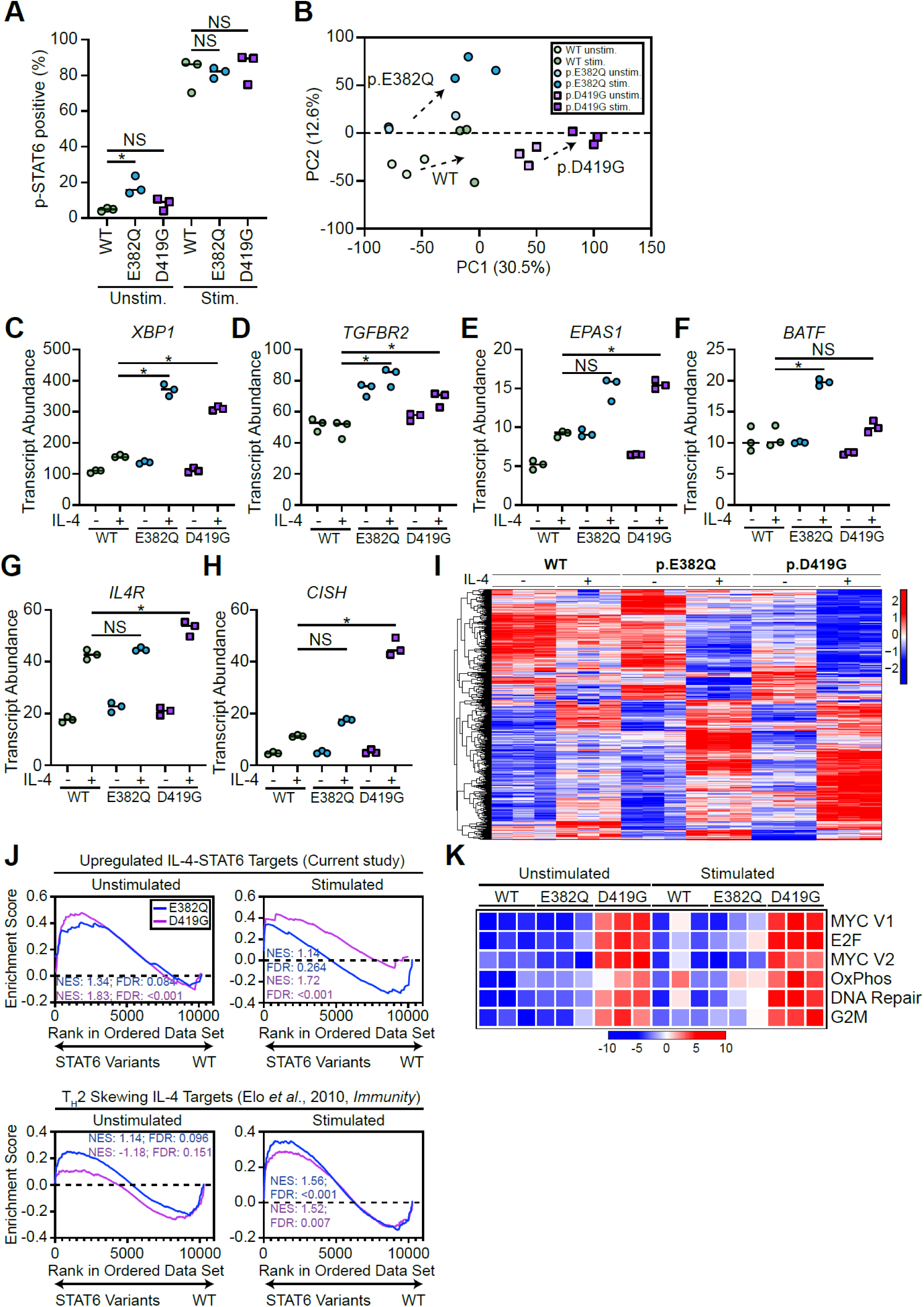
p.E382Q and p.D419G *STAT6* variants lead to increased STAT6 activity in Jurkat T cells. **A)** Quantification of phospho-STAT6-positive WT-, p.E382Q-, or p.D419G-transduced Jurkat T cells before and after treatment with IL-4 (100ng/mL for 1h), n=3. **B)** Principal component analysis (PCA) comparing unstimulated and stimulated (100 ng/mL IL-4 for 4 h) WT (green), p.E382Q (blue), p.D419G (purple) *STAT6* transduced Jurkat T cells. PC1 and PC2 contribution is shown in brackets. **C-H)** Normalized counts comparing stimulated WT (green) vs. p.E382Q (blue) or p.D419G (purple), for **C)** *XBP1*, **D)** *TGFBR2*, **E)** *EPAS1*, **F)** *BATF*, **G)** *IL4R*, **H)** *CISH*. **I)** Heatmap representation of normalized counts of a transcription set defined as IL-4 targets in each STAT6 Jurkat T cells. *adjusted P-val<0.05. **J)** GSEA plots for curated STAT6 and IL-4-T_H_2 targets genes, comparing WT vs. either p.E382Q (blue) or p.D419G (purple) at both baseline and after stimulation with IL-4. Normalized enrichment score (NES) and adjusted p-value are shown. **K)** Sample level enrichment analyses (SLEA) of significantly enriched immune pathways from MSigDB Hallmark in unstimulated and IL-4-stimulated samples, comparing WT vs either p.E382Q or p.D419G. Heatmap is normalized across the rows and shown as relative expression.

Transcriptomically, WT-, p.E382Q-, and p.D419G-transduced cells clustered separately from each other both at baseline and after stimulation with IL-4 (**Fig 3B**). This confirmed that all variant cells were responsive to IL-4, and identified a unique transcriptomic signature for each. Differential gene expression analysis revealed significantly increased transcript abundance of known STAT6 target genes, including *IL4R*^*52*^, *CISH*^*45*^ and *EPAS*^*42*^, and genes that have been implicated in atopic disease, including *XBP1*^*53*^ and *TGFBR2*^*54*^ in p.E382Q and p.D419G transduced cells when compared to WT transduced controls (**Fig 3C-H**).

In response to IL-4 stimulation, we observed 54, 145, and 159 significantly upregulated genes in WT-, p.E382Q-, and p.D419G-transduced Jurkat T cells, respectively (**Fig 3I, Supp Fig 2A**). Interestingly, p.E382Q and p.D419G had 67 and 80 uniquely increased hits, which did not overlap with WT, nor with each other. On the other hand, p.E382Q and p.D419G had 108 and 54 uniquely downregulated genes (**Fig 3I, Supp Fig 2A**). This suggests that the altered activity of both p.E382Q and p.D419G is not restricted to enhanced activity on known STAT6 targets alone.

To further investigate the differential gene expression patterns, we performed gene set enrichment analyses (GSEA). We first defined IL-4-STAT6 targets as genes that were significantly upregulated in stimulated WT STAT6-transduced Jurkat T cells (**Supp Table 2**). We found enrichment of these targets in both p.E382Q and p.D419G Jurkats only at baseline (p.E382Q NES 1.34, adj-pvalue 0.084; p.D419G NES 1.83, adj-pvalue <0.001), but not after IL-4 stimulation. Notably, after IL-4 stimulation both p.E382Q and p.D419G STAT6 Jurkats showed significantly increased enrichment of IL-4 responsive T_H_2 skewing targets^42^ (p.E382Q NES 1.56, adj-pvalue <0.001; p.D419G NES 1.52, adj-pvalue 0.007)(**Fig 3J**).

GSEA using the MSigDB Hallmark gene sets showed no significant difference in enrichment in immunological pathways between WT and p.E382Q STAT6 both at baseline and in response to stimulation (**Fig 3K, Supp Table 3**). However, p.D419G STAT6 exhibited significantly increased enrichment in proliferation (Myc, G2M, DNA repair, E2F) pathways, consistent with the fact that p.D419G (but not p.E382Q) is reported as a recurrent somatic mutation in lymphoma^45,55,56^, establishing this variant as oncogenic.

### JAK inhibitors reduce enhanced STAT6 phosphorylation induced by p.E382Q and p.D419G STAT6 in HEK293 cells

Having demonstrated that both p.E382Q and p.D419G STAT6 are GOF variants, we tested whether small molecule inhibitors that target the STAT6 pathway could potentially be clinically beneficial. We selected JAK inhibitors, ruxolitinib and tofacitinib, as both of these drugs are already used clinically for treating atopic disease^57,58^. Both inhibitors effectively decreased the enhanced STAT6 phosphorylation observed at baseline and after IL-4 stimulation in cells expressing the p.E382Q or p.D419G STAT6 variants (**Fig 4A-B**). This suggests that JAK inhibitors may be promising treatment options for patients with GOF *STAT6* variants.

**Figure 4:**
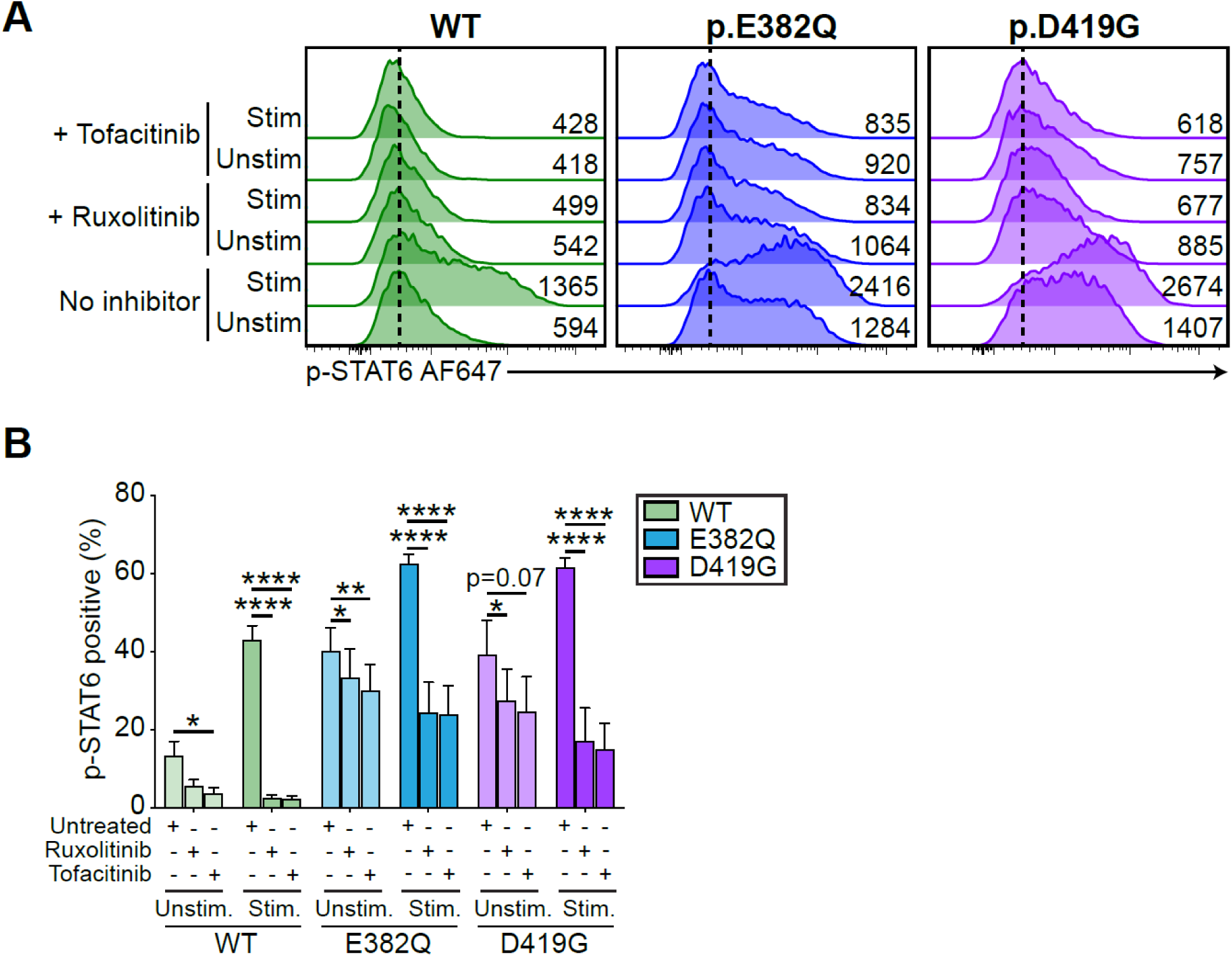
JAK inhibitor treatment decreases gain-of-function STAT6 activity caused by p.E382Q and p.D419G STAT6. **A)** phospho-STAT6 expression in transfected HEK293 cells pre-treated with ruxolitinib (10μM 2hr) or tofacitinib (4μM 2hr), before and after stimulation with IL-4 (100ng/mL 1h). **B)** Quantification of A), n=3. One-way ANOVA and Tukey’s post-hoc test. *p<0.05, **p<0.01, ***p<0.001, ****p<0.0001.

## DISCUSSION

In this study, we present a combination of clinical, molecular, and transcriptional evidence of a new human disorder caused by germline autosomal dominant GOF *STAT6* variants in two patients with life-long severe allergic disease. These variants led to increased STAT6 phosphorylation, increased STAT6 target gene expression, and T_H_2 skewed transcriptional profile. Although the full phenotype of GOF *STAT6* variants will only be uncovered through the identification of additional affected individuals, we propose to classify human germline autosomal dominant GOF STAT6 syndrome as a novel primary atopic disorder^24-26^. Based on our study, possible clinical ‘red flags’ for this new disorder include: (i) early life onset; (ii) peripheral blood eosinophilia, (iii) elevated serum IgE, (iii) widespread, treatment-resistant atopic dermatitis, (iv) multiple food and drug allergies; (v) recurrent skin and respiratory infections; (vi) eosinophilic gastrointestinal disorder, including eosinophilic esophagitis; (vii) allergic rhinoconjunctivitis; and (viii) vascular malformations of the brain.

STAT6 is intimately linked to the biology of allergic inflammation. The central and most studied role of STAT6 is in mediating the biological effects of IL-4, a cytokine necessary for T_H_2 differentiation, B cell survival, proliferation and class switching to IgE^9,42^, and driving M2 macrophage polarization^59^. In T cells, STAT6 activation induces the expression of GATA3, the master regulator of T_H_2 differentiation, which in turn enhances expression of IL-4, IL-5 and IL-13, cytokines necessary for promoting allergic responses by activating mast cells and eosinophils^60^. Presence of greater T_H_2 cell populations, or T_H_2 cells producing copious amounts of IL-4/IL-5/IL-13, could be a driver of the observed allergic phenotype presented in our patients. Elevated IgE in partnership with mast cells is important for both acute and chronic manifestations of allergic disorders and can be an additional driver of the allergic diathesis^61^. STAT6 hyperactivation in airway epithelial cells and resident dendritic cells can further create an environment favouring asthma and chronic lung disease, as this would induce production of chemokines that promote T_H_2 cells and eosinophil recruitment^62,63^. Population genetics provide further support for the central role that STAT6 plays in the development of human allergic disease. Multiple independent genome wide association studies (GWAS) have found that polymorphisms in *STAT6* associate with many allergic conditions (**Table 2**). Our study further expands this appreciation of the role of STAT6 in human disease by establishing that heterozygous GOF variants cause a monogenic form of severe allergic disease.

**Table 2.** Number of published genome-wide association (GWAS) studies linking polymorphisms (SNPs) in *STAT6* to common allergic diseases in the population. Significant genome-wide associations (P < 5×10^−8^) between *STAT6* SNPs and all relevant allergic traits were obtained through NHGRI-EBI GWAS Catalog (ebi.ac.uk/gwas/).

| Phenotype | Number of published associations | References |
| --- | --- | --- |
| Asthma* | 13 | 76-88 |
| Eosinophil count | 6 | 76,89-93 |
| Allergic disease | 3 | 80,94,95 |
| Atopic dermatitis/Eczema | 3 | 81,93,96 |
| Serum IgE level | 2 | 97,98 |
| Allergic sensitization | 2 | 99,100 |
| Allergic rhinitis | 1 | 81 |
| Eosinophilic gastrointestinal disorders | 1 | 101 |

\*Includes asthma, childhood-onset asthma, adult-onset asthma, and atopic asthma

The fatal cerebral aneurysm in P1 (p.E382Q) was not clinically anticipated, but it is possible that the *STAT6* GOF variant also increased the risk of developing cerebral aneurysms. Intracranial aneurysms have been reported in patients with both STAT3 LOF and STAT1 GOF^64-66^. Increased activation of other STAT family members, including STAT2, STAT3 and STAT5 have also been observed in human abdominal aortic aneurysms (STAT6 was not evaluated), although it is not clear whether enhanced STAT phosphorylation contributes to aneurysms or is the result of inflammation caused by aneurysms^67^. As more individuals with *STAT6* GOF variants are identified, the possibility of cerebral vascular anomalies warrants investigation.

A GOF STAT6 model (designated STAT6VT) has previously been described *in vitro*^68^ and has been used to study chronic atopic dermatitis in mouse models^69,70^. STAT6VT has the substitution of two amino acid residues, at positions 547 and 548, in the SH2 domain resulting in a STAT6 mutant that is constitutively active in an IL-4 independent manner and is unresponsive to IL-4 stimulation^68^. STAT6VT differs from the variants we describe as the latter are in the DNA binding domain (**Fig 1**) and do respond to IL-4 stimulation (**Fig 2-4**). Nevertheless, the humans we identified with STAT6 GOF variants and STAT6VT mice share a number of key features of the allergic diathesis, including elevated serum IgE and chronic atopic dermatitis.

There is now a growing list of human single gene defects that cause the classic hyper-IgE phenotypic triad of eczema, recurrent skin and lung infections, and elevated serum IgE^26,71^. Autosomal dominant hyper-IgE syndrome caused by dominant negative variants in *STAT3* (aka. Job’s syndrome or STAT3 LOF) is the best characterized of these conditions, but this list of disorders does include both autosomal dominant (*STAT3, CARD11, ERBIN*) and autosomal recessive (*DOCK8, PGM3, IL6ST*) conditions^72,73^. Notably, the patients we identified with STAT6 GOF variants did have some of the extra-immunological features typical of STAT3 LOF (i.e. hyperextensible joints, fractures, vascular anomalies^73^).

Based on the findings reported in this study, we suggest that heterozygous GOF variants in STAT6 be added to the list of autosomal dominant causes of the hyper-IgE phenotype. While each of the conditions known to cause a hyper-IgE phenotype has some specific clinical features (e.g. viral skin infections are a defining feature of DOCK8 deficiency^74,75^), there is considerable clinical overlap and clinically-approved testing of these pathways is rarely available. Therefore, we recommend genetic testing as the most efficient initial diagnostic approach to patients who experience severe allergic disease beginning very early in life. Finally, in common with other primary atopic disorders^31^, we demonstrate that JAK inhibition may be an effective treatment for patients with GOF *STAT6* variants.

While this study has many strengths, notably the extreme allergic phenotype of the two patients combined with in-depth functional assessment of their *de novo STAT6* variants, a limitation is our lack of validation in primary patient cells due to the death of P1 and challenges with phlebotomy in P2 due to the severity of her atopic dermatitis. Despite this limitation, our study does identify GOF variants in *STAT6* as a novel monogenic allergic disorder. We anticipate that this discovery will facilitate the recognition of more affected individuals and, with time, a full definition of the human phenotype caused by germline human *STAT6* GOF variants will emerge.

## Data Availability

All data produced in the present study are available upon reasonable request to the authors.

## Acknowledgements

This work was supported by grants from the Canadian Institutes of Health Research (PJT 178054) (S.E.T.), Genome British Columbia (SIP007) (S.E.T.), and BC Children’s Hospital Foundation. S.E.T. holds a Tier 1 Canada Research Chair in Pediatric Precision Health and the Aubrey J. Tingle Professor of Pediatric Immunology. M.S. is supported by a CIHR Frederick Banting and Charles Best Canada Graduate Scholarships Doctoral Award (CGS-D) and University of British Columbia Four Year Doctoral Fellowship (4YF). H.Y.L. is supported by a CGS-D, 4YF, Killam Doctoral Scholarship, Friedman Award for Scholars in Health, and a BC Children’s Hospital Research Institute Graduate Studentship. M.V.S. is funded by the Vanier Canada Graduate Scholarship and the University of British Columbia 4-Year Doctoral Fellowship (4YF). We would also like to acknowledge the Biomedical Research Centre Sequencing Core (BRC-Seq) for their assistance with RNA-Sequencing and processing.

## Contribution

Me.S., G.B.R, H.M.B, Mi.S, M.L.M, H.A, K.L.D., and S.E.T enrolled patients and analyzed clinical data; J.J.L, A.M, W.W.W., C.D.M.v.K conducted genetic screening and identified variants of significance. M.S., O.F, R.v.d.L, and P.A.R performed, structural modeling, bioinformatic and statistical analysis; M.S., H.Y.L., M.V.S, K.L.D, S.L., and J.D. performed the laboratory experiments; M.S., H.Y.L., M.V.S, and S.E.T. wrote and edited the manuscript. All authors reviewed the final manuscript.

## SUPPLEMENTAL FIGURES

**Supplementary Figure 1:**
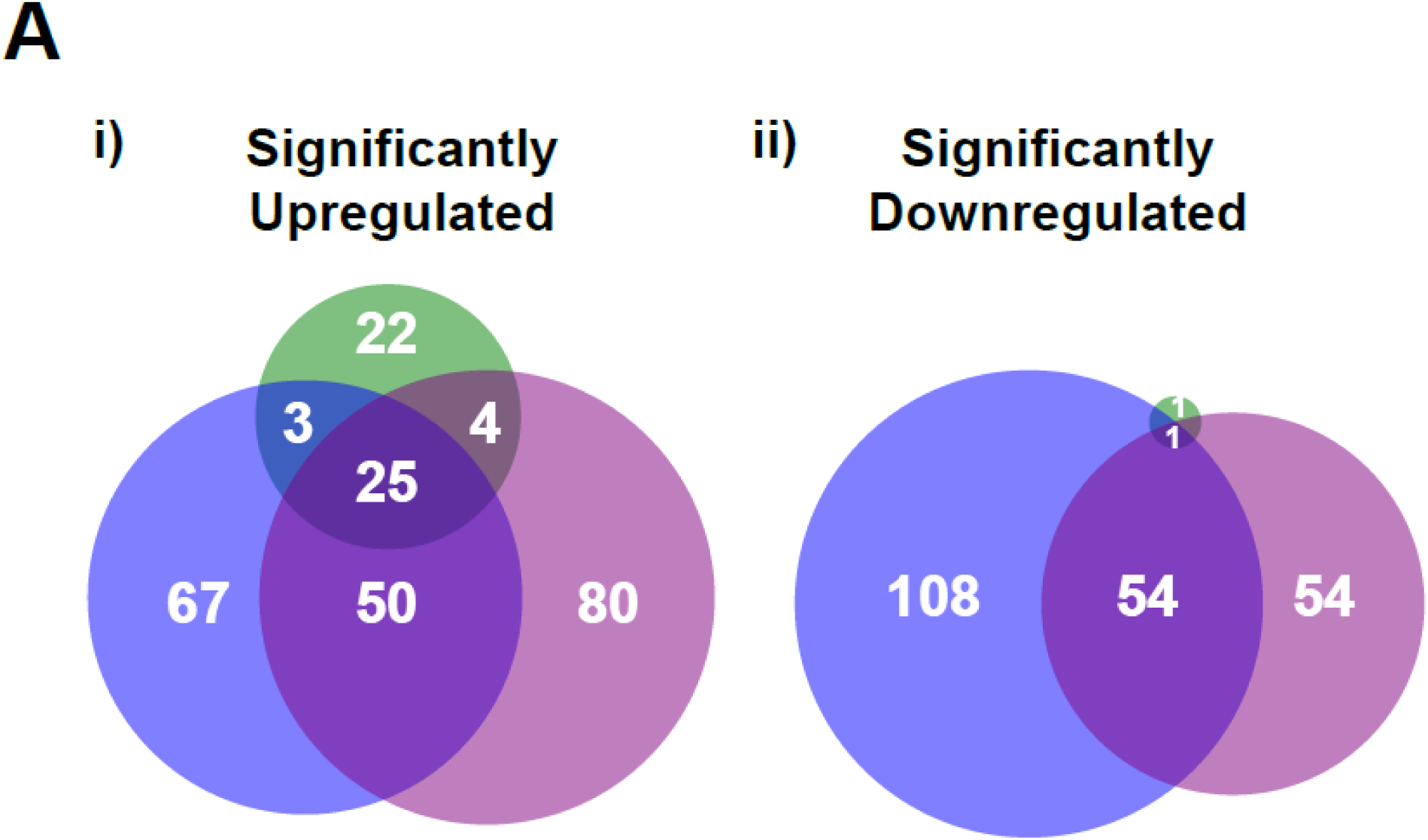
Extended data on STAT6 variant-transduced Jurkat T cells. **A)** Significantly upregulated (i) and downregulated (ii) genes upon IL-4 treatment in WT (green), p.E382Q (blue) and p.D419G (purple) in Jurkat cells as shown via Venn diagram.

## SUPPLEMENTAL TABLES

**Supplementary Table 1:** Variant annotation and pathogenicity prediction of the variants reported in the two patients.

| Variant Annotation |  |  |
| --- | --- | --- |
|  | P1 | P2 |
| Chromosome | 12 | 12 |
| Genomic Position (GRCh37) | 57498315 | 57496661 |
| cDNA Position (NM_001178079.2) | 1144 | 1256 |
| Nucleotide Reference | G | A |
| Nucleotide Variant | C | G |
| Protein Variant (NP_001171550.1) | p.Glu382Gln (p.E382Q) | p.Asp419Gly (p.D419G) |
| Zygosity | Heterozygous | Heterozygous |
| Inheritance | *de novo* | *de novo* |
| dbSNP153 | No entry | No entry |
| gnomAD (v3.1.1) | No entry | No entry |
| COSMIC (v95) | Somatic not reported | Somatic reported, 26 entries (COSV55668829) |
| *in silico* Pathogenicity Prediction Models |  |  |
| CADD (v1.6) | 27.7 | 29.3 |
| SIFT | Deleterious (0.026) | Deleterious (0.001) |
| PolyPhen-2 | Probably Damaging (1) | Probably Damaging (1) |
| LRT | Deleterious (0) | Deleterious (0) |
| MutationTaster | Disease Causing (1) | Disease Causing (1) |
| PROVEAN | Damaging (-2.71) | Damaging (-4.83) |
| MetaSVM | Tolerable (-0.538) | Damaging (0.610) |
| M-CAP | Possibly pathogenic (0.106) | Possibly pathogenic (0.294) |
| FATHMM MKL Coding | Deleterious (0.986) | Deleterious (0.701) |

**Supplementary Table 2:** IL-4/STAT6 target genes in Jurkat. IL-4/STAT6 target genes are shown that were either upregulated (green) or downregulated (red) in WT-STAT6 transduced Jurkat after stimulation with IL-4 (100ng/mL) for 4 hours.

| IL4-STAT6 targets in Jurkat |  |  |  |
| --- | --- | --- | --- |
| *SOCS1* | *STK17B* | *IRF2BP2* | *IDI1* |
| *MAL* | *SLC39A8* | *UBE2B* | *CHST2* |
| *FAM171A1* | *MZT1* | *TEX30* | *RPF2* |
| *CD244* | *MBNL1* | *ABCD3* | *LYPLA1* |
| *BCL6* | *ETS2* | *SCOC* | *CCNC* |
| *IL4R* | *PMAIP1* | *NDUFA5* | *CENPH* |
| *CISH* | *ENDOD1* | *SNX10* | *CBX3* |
| *RAB11FIP1* | *TMED7* | *LZTFL1* | *HINT3* |
| *TREML2* | *ST8SIA4* | *COX20* | *XIAP* |
| *SNTB1* | *PRKCQ-AS1* | *GRK6* | *CD164* |
| *PECAM1* | *TSNAX* | *LINC00493* | *STK17A* |
| *GCSAM* | *NDUFB5* | *DCK* | *DSTN* |
| *RASGRP1* | *XBP1* | *STAMBPL1* | *ARPC5* |
| *GATSL3* | *RFK* | *SGCB* | *RAP1GAP* |
| *FAM120AOS* | *TESPA1* | *DENR* | *CLSTN3* |

**Supplementary Table 3:**
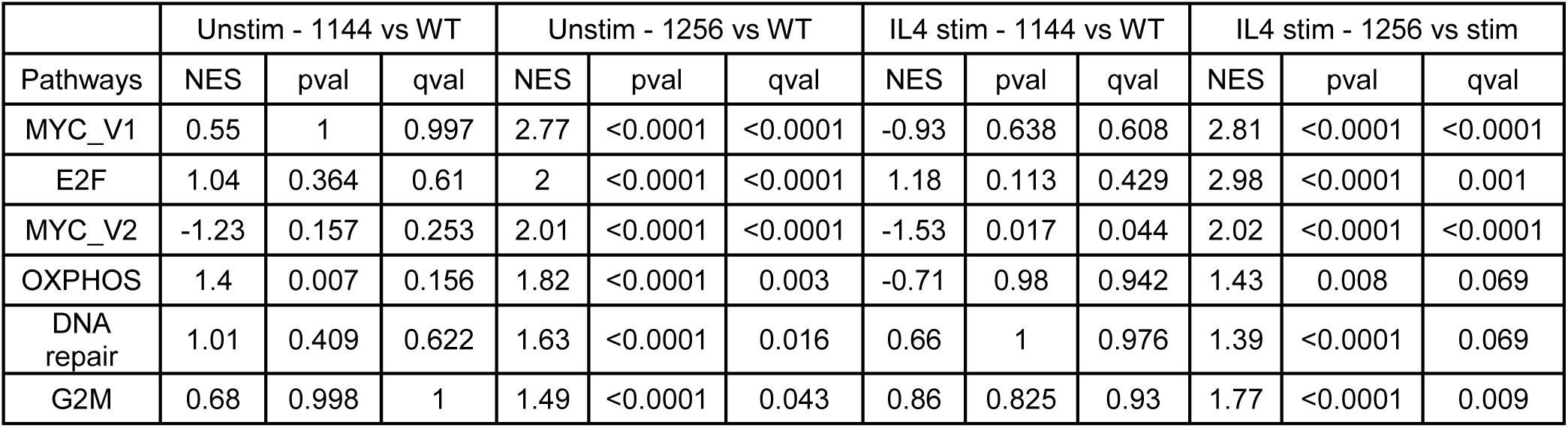
Statistics for enrichment analysis done on Jurkat T cells, comparing WT-STAT6 with either p.E382Q or p.D419G at baseline or after stimulation.

